# Potential roles of social distancing in mitigating the spread of coronavirus disease 2019 (COVID-19) in South Korea

**DOI:** 10.1101/2020.03.27.20045815

**Authors:** Sang Woo Park, Kaiyuan Sun, Cécile Viboud, Bryan T. Grenfell, Jonathan Dushoff

## Abstract

On January 20, 2020, the first COVID-19 case was confirmed in South Korea. After a rapid outbreak, the number of incident cases has been consistently decreasing since early March; this decrease has been widely attributed to its intensive testing. We report here on the likely role of social distancing in reducing transmission in South Korea. Our analysis suggests that transmission may still be persisting in some regions.

## 1 Introduction

Since its first appearance in Wuhan, China, in December 2019 [1], coronavirus disease (COVID-19) has spread internationally, including to South Korea. The first COVID-19 case in South Korea was confirmed on January 20, 2020, from a traveling resident of Wuhan, China [2]. In February, the disease spread rapidly within a church community in the city of Daegu [2]. The chains of transmission that began from this cluster distinguish the epidemic in South Korea from that in any other countries: As of March 24, 2020, 9,037 cases were confirmed, of which 56% were related to the church and 27% were in their 20s [2]. South Korea’s intensive testing using novel contact tracing techniques allowed rapid identification and isolation cases and reduction of onward transmission [3, 4, 5]. Here, we describe potential roles of social distancing in mitigating the spread of COVID-19 in South Korea by using metro traffic data to compare epidemics in two major cities.

## 2 Materials and Methods

### 2.1 Data description

We analyzed epidemiological data describing the COVID-19 outbreak in South Korea between January 20–March 16, 2020. Daily number of reported cases in each geographic region was transcribed from press releases by the Korea Centers for Disease Control and Prevention (KCDC) [2]. Partial line lists were translated and transcribed from press releases by the KCDC and various local and provincial governments [6, 7, 8, 9, 10, 11, 12]. All data and code are stored in a publicly available GitHub repository: https://github.com/parksw3/Korea-analysis.

We compared epidemiological dynamics of COVID-19 from two cities in which the largest number of COVID-19 cases have been reported: Daegu and Seoul. Between January 20– March 16, 2020, 6,083 cases from Daegu and 248 from Seoul were reported by the KCDC. The epidemic in Daegu is characterized by a single, large peak followed by a gradual decrease, whereas the epidemic in Seoul consists of several small outbreaks (Fig. 1).

**Figure 1:**
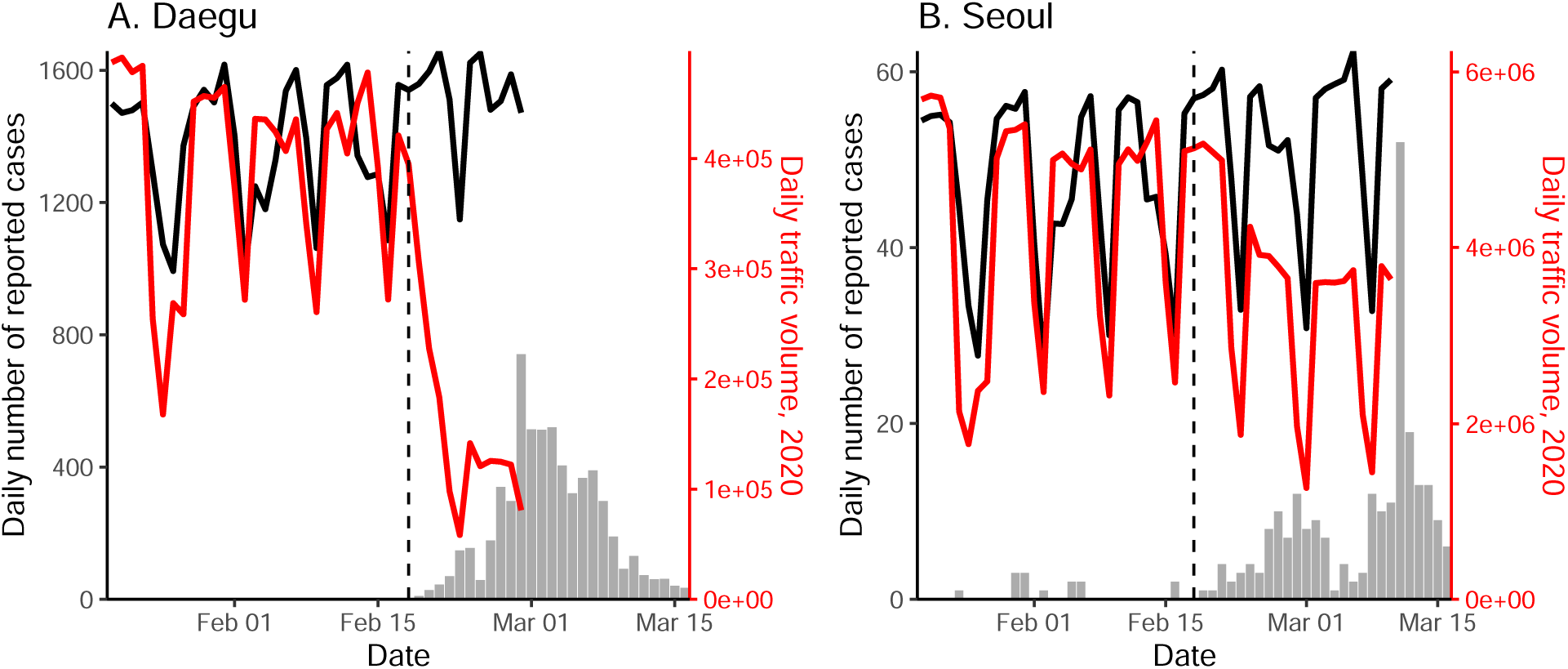
Comparison of epidemiological and traffic data from Daegu and Seoul. Solid lines represent the daily metro traffic volume in 2020 (red) and mean daily metro traffic volume between 2017–2019 (black). Daily traffic from previous years have been shifted by 1–3 days to align day of the weeks. Vertical lines indicate Feb 18, 2020, when the first case was confirmed in Daegu.

Daily metro traffic in Daegu and Seoul between 2017–2020 was obtained from `data.go.kr` and `data.seoul.go.kr`, respectively. We tabulated the total number of individuals who accessed the subway or monorail (using Seoul lines 1–9, and Daegu lines 1–3; Fig. 1). Soon after the first church-related case was confirmed in Daegu on Feb 18, 2020, the daily traffic volume decreased by about 80% and 50% in Daegu and Seoul, respectively.

### 2.2 Time-dependent reproduction number

To estimate the time-dependent reproduction number *R*_*t*_ (the average number of secondary cases caused by an average individual, given conditions at time *t* [13]), we first estimated daily incidence of infection from the daily number of reported cases by the KCDC [2]. We adjusted the number of reported cases to account for changes in testing criteria, which occurred 4 times between January 20–March 16, 2020. Then, we inferred onset-to-confirmation delay distributions from the partial line list and combined them with previously estimated incubation period distribution (Table 1) to obtain probability distributions for date of infection for each reported case. We accounted for right-censoring by dividing the daily incidence by the probability that a case infected on a given day would have been reported before March 16, 2020. Implementation details are provided in the Supplementary Materials.

**Table 1:** Assumed incubation and generation-interval distributions. Gamma distributions are parameterized using its mean and shape. Negative binomial distributions are parameterized using its mean and dispersion. Priors are chosen such that the 95% quantiles of prior means and standard deviations are consistent with previous estimates.

|  | Parameterization | Priors | Source |
| --- | --- | --- | --- |
| Incubation period distribution | $\text{Gamma}(\mu_I, \mu_I^2/\sigma^2)$ | $\mu_I \sim \text{Gamma}(6.5 \text{ days}, 145)$<br>$\sigma \sim \text{Gamma}(2.6, 25)$ | [14] |
| Generation-interval distribution | $\text{NegativeBinomial}(\mu_G, \theta)$ | $\mu_G \sim \text{Gamma}(5 \text{ days}, 62)$<br>$\theta \sim \text{Gamma}(5, 20)$ | [15, 16] |

We estimated the time-dependent reproduction number using the renewal equation with a 14-day sliding window [13]:

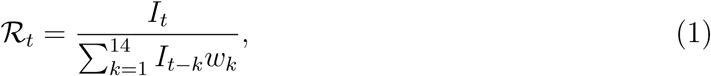

where *I*_*t*_ is the reconstructed incidence time series (i.e., the number of infected cases on day *t*) and *w*_*k*_ is the generation-interval distribution randomly drawn from a prior distribution (Table 1). We weighted each sample of ℛ_*t*_ by a gamma probability distribution with a mean of 2.6 and a standard deviation of 2 to reflect prior knowledge [17] and took weighted quantiles to calculate the medians and associated 95% credible intervals. We estimated ℛ_*t*_ between February 2, 2020 (14 days after the first confirmed case was imported) and March 10, 2020 (after this point the effects of censoring are too strong for reliable estimates).

## 3 Results

Fig. 2 compares the reconstructed incidence (A,B) and estimates of ℛ_*t*_ (C,D) in Daegu and Seoul. In Daegu, incidence peaked shortly after the first case was confirmed and then decreased (Fig. 2A). Likewise, the estimates of ℛ_*t*_ gradually decrease and eventually drop below 1 about a week after the reporting of its first case, coinciding with the decrease in the metro traffic volume (Fig. 2C). The initial decrease in ℛ_*t*_ may reflect behavior change within the church; the first confirmed case in Daegu became symptomatic on February 7, 2020, and visited the church on February 9 and 16, 2020 [2]. Our estimates of ℛ_*t*_ for Daegu are consistent with the estimates of ℛ_*t*_ for South Korea by Abbott *et al*. [17] — their estimates drop below 1 slightly later because they rely on number of symptomatic cases instead.

**Figure 2:**
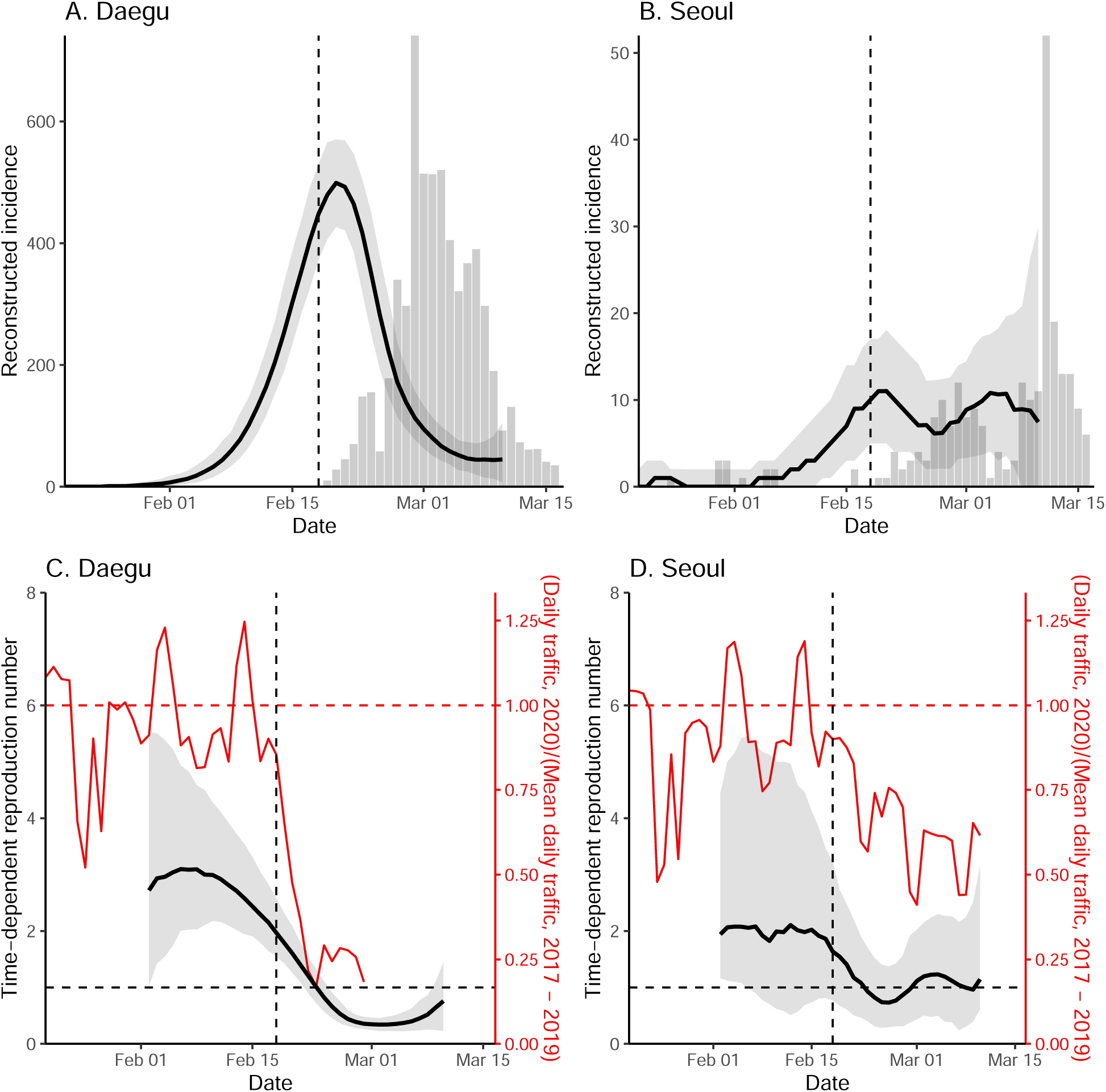
Comparison of reconstructed incidence and time-dependent reproduction number in Daegu and Seoul. Black lines and gray ribbons represent the median estimates of reconstructed incidence (A,B) and ℛ_*t*_ (C,D) and their corresponding 95% credible intervals. Bar plots show the number of reported cases. Red lines represent the normalized traffic volume. Vertical lines indicate Feb 18, 2020, when the first case was confirmed in Daegu.

In Seoul, estimates of ℛ_*t*_ decrease slightly but remain around 1 (Fig. 2D). Our analysis suggests that social distancing in Seoul was less intense, and this could be why reduction in spread was less sharp. Stronger distancing or further intervention will be necessary to reduce ℛ_*t*_ below 1.

While we find clear, positive correlations between the normalized traffic and the median estimates of ℛ_*t*_ in both Daegu (*r* = 0.90; 95% CI: 0.79–0.95) and Seoul (*r* = 0.76; 95% CI: 0.59–0.87), these correlations are conflated by time trends, and are also likely conflated by other measures that could have affected ℛ_*t*_. We do not find clear signatures of lagged correlation between ℛ_*t*_ and traffic volume (Supplementary Materials). Similar patterns in the estimates of _*t*_ are found in directly surrounding provinces (Gyeongsangbuk-do and Gyeonggi-do), providing support for the robustness of our analysis (Supplementary Materials).

## 4 Discussion

The South Korean experience with COVID-19 provides evidence that epidemics can be suppressed with less extreme measures than those taken by China [18]. It demonstrates the necessity of prompt identification and isolation of cases in preventing further spread [3, 4, 5]. Our analysis reveals potential roles of social distancing in mitigating the COVID-19 epidemic in South Korea. Even though social distancing alone may not be able to fully prevent the spread of the disease, its ability to flatten the epidemic curve (cf. Fig. 2B,D) reduces burden for healthcare system and provides time to plan for the future [19].

Our study is not without limitations. We did not account for differences in the delay distributions or changes in the number of tests among cities. The intensity of intervention is likely to vary across regions given that majority of COVID-19 cases in South Korea were reported from Daegu. We did not have sufficient data to account for these factors. Nonetheless, the robustness of our findings is supported by the sensitivity analyses (Supplementary Materials). We were also unable to distinguish local and imported cases, which may overestimate ℛ_*t*_ [20]. We were able to perform a separate analysis for Seoul that accounts for imported cases using line list provided by the Seoul Metropolitan Government; our qualitative conclusions remained robust (Supplementary Materials).

Our analysis focused on comparing metro traffic, which serves as a proxy for the degree of social distancing, with epidemiological dynamics in two cities. The 80% decrease in traffic volume suggests that the strength of social distancing in Daegu may be comparable to that in Wuhan, China [21]. However, we are not able to directly estimate the effect of social distancing on epidemiological dynamics. Other measures, such as intensive testing of core transmission groups and school closure, are also likely to have affected the changes in ℛ_*t*_ [2]. Future studies should consider quantifying contributions of different measures in preventing the spread.

Finally, our study highlights the importance of considering geographical heterogeneity in estimating epidemic potential. The recent decrease in the number of reported cases in South Korea is driven by the sharp decrease in Daegu. Our analysis reveals that the epidemic may still persist in other regions, including Seoul and Gyeonggi-do; recent reports from Seoul and Gyeonggi-do (around 10 new cases almost every day between March 11–24, 2020) provide further support for our conclusion [2]. Unless the reproduction number can be reduced below 1 in all regions, small outbreaks may continue to occur in South Korea.

## Contribution

Data collection: SWP; conceptualization: SWP, CV; analysis: SWP; first draft: SWP, JD. All authors contributed to the writing and approval of the final report.

## Data Availability

All data and code are stored in a publicly available GitHub repository: https://github.com/parksw3/Korea-analysis.

## Supplementary Materials

### Epidemiological data

The daily number of reported cases from each region was translated and transcribed from the KCDC press release [2]. Following the KCDC’s protocol, the daily number of reported cases prior to February 20, 2020, reflects the number of confirmed cases on each day. Between February 21 – March 1, 2020, the daily number of reported cases reflects the number of reported cases within the last 24 hours (9 AM to 9 AM). On March 2, 2020, the daily number of reported cases reflects the number of cases that were reported between 9 AM March 1, 2020, and 12 AM March 2, 2020. Since then, the daily number of reported cases reflects the number of reported cases within the last 24 hours (12 AM to 12 AM). The number of negative cases was not reported on January 25 and 31, 2020; we took the average of cumulative negative cases from one day before and after these dates instead to impute missing values. The daily number of reported cases by the KCDC may be slightly different from the reports by each city’s government as some cases may be transferred after they are confirmed. The sum of daily number of reported cases by the KCDC may be also slightly different from the cumulative number of cases reported the KCDC because it does not reflect possible location changes of the confirmed cases after reporting.

### Reconstruction of incidence time series

Testing criteria expanded 4 times between January 20–March 16, 2020: January 28, February 7, February 20, and March 2, 2020. We accounted for these changes by assuming that the proportion positive should remain roughly constant if we follow a consistent protocol of identifying and deciding whom to test. To do so, we calculated the relative proportion of positive cases during each period (divided by the the between-period mean) and multiplied the daily number of reported cases by the relative proportions of the corresponding criterion. Sensitivity analyses showed that results are robust to these adjustments.

We then estimated time-dependent *backward* onset-to-confirmation delay distributions from the partial line list: Given a cohort of infected individuals who were confirmed on the same day, what is the probability distribution of the onset-to-confirmation delay? The backward delay distribution depends on changes in the number of symptomatic cases — e.g., when the number of symptomatic cases are increasing, the backward delay distribution is likely to be shorter because individuals are more likely to have developed symptoms recently. The backward delay distribution was inferred using a negative-binomial regression with log-link using the `brms` package [22]. Time-dependent mean of the negative binomial distribution is modeled using splines. We assumed weakly informative priors on the fixed effects: normal distributions with mean of 0 and standard deviation of 2; note that these distributions are priors on link scale.

For each posterior sample of the backward delay distribution, we drew a random sample of onset-to-confirmation delay and incubation period for each confirmed case. This allowed us to obtain posterior samples of possible infection dates for each case, which were then converted into posterior samples of incidence time series.

To account for right-censoring in the reported cases, we also estimated time-dependent *forward* onset-to-confirmation delay distribution using the same negative-binomial regression model: Given a cohort of infected individuals who became symptomatic on the same day, what is the probability distribution of the onset-to-confirmation delay? The forward delay distribution reflects the changes in the accuracy of case identification — e.g., a decrease in the delay reflects improvement in accuracy.

To estimate the forward delay distribution, we modified the stan code from the negative-binomial regression that we used to infer the backward delay distribution to account for right-censoring (in the observed delays) and ran the code using the `rstan` package [23]. In particular, we modified the likelihood of the negative-binomial regression such that given a delay of *x*_*i*_ days, symptom onset day *t*_*i*_ and the day of measurement of *t*_max_, the likelihood of observing the delay is given by:

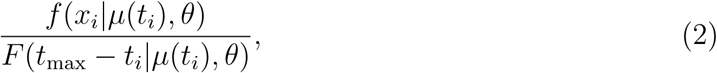

where *f* is the negative binomial distribution with time-dependent mean *µ*(*t*_*i*_) and dispersion parameter *θ*. This likelihood accounts for the fact that the delay between symptom onset and confirmation cannot be longer than *t*_max_ − *t*_*i*_ (otherwise, the case will be reported after *t*_max_). Convergence is assessed by the lack of warning messages from the `rstan` package [23]. For each combination of date of infection and a posterior sample of the forward delay distribution, we drew 1000 samples of incubation periods and onset-to-confirmation delays and calculated the median probability that an individual infected on a given day will be confirmed before March 16, 2020. Finally, we divided the daily number of infected cases by the median probability this probability. We used the reconstructed incidence time series to estimate ℛ_*t*_.

**Figure S1:**
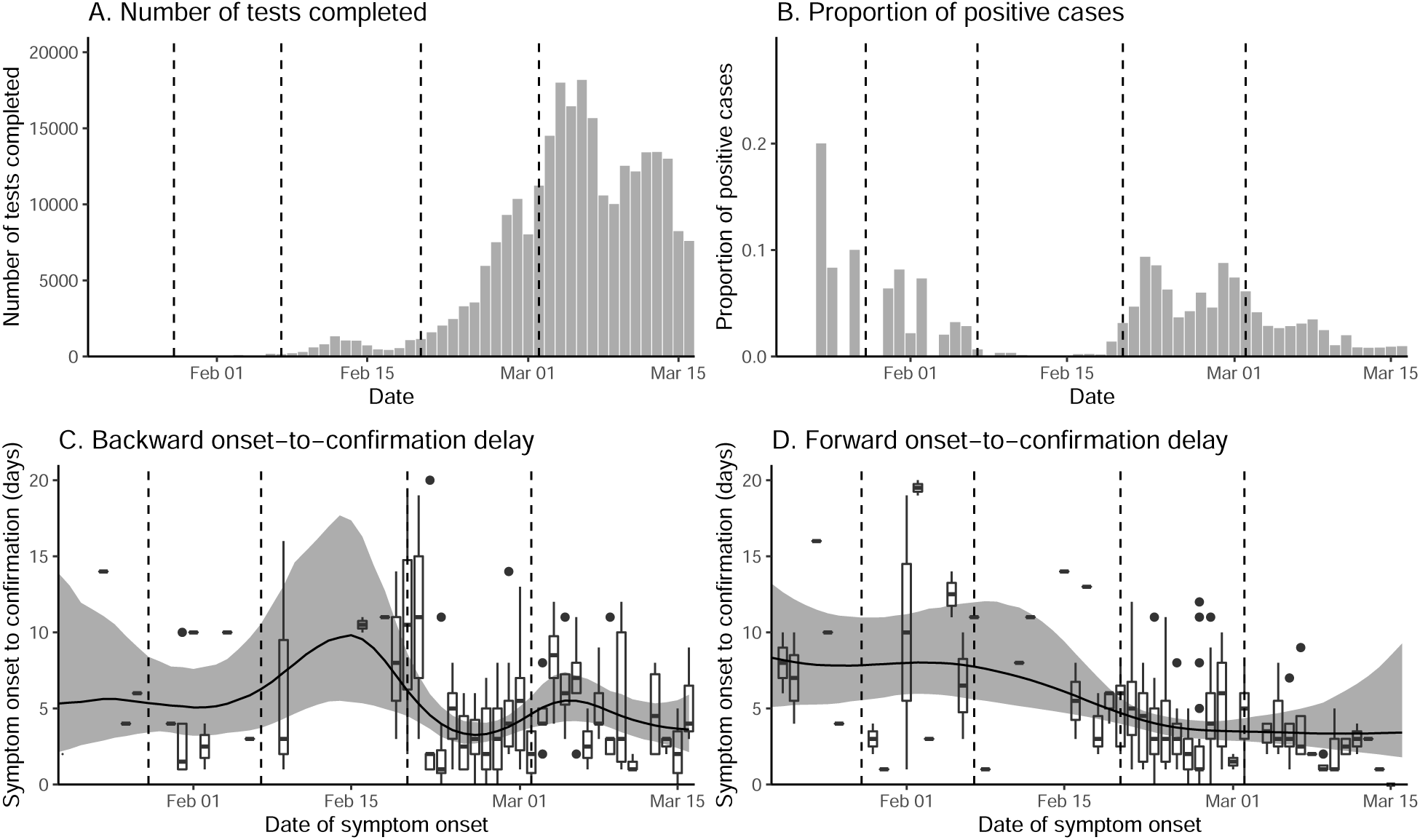
Changes in the number of tests and delay distributions over time. Vertical lines indicate the date on which testing criteria expanded. Box plots (C–D) represent the observed delays. Black lines and gray ribbons represent the median estimates of the mean delays and their associated 95% credible intervals.

**Figure S2:**
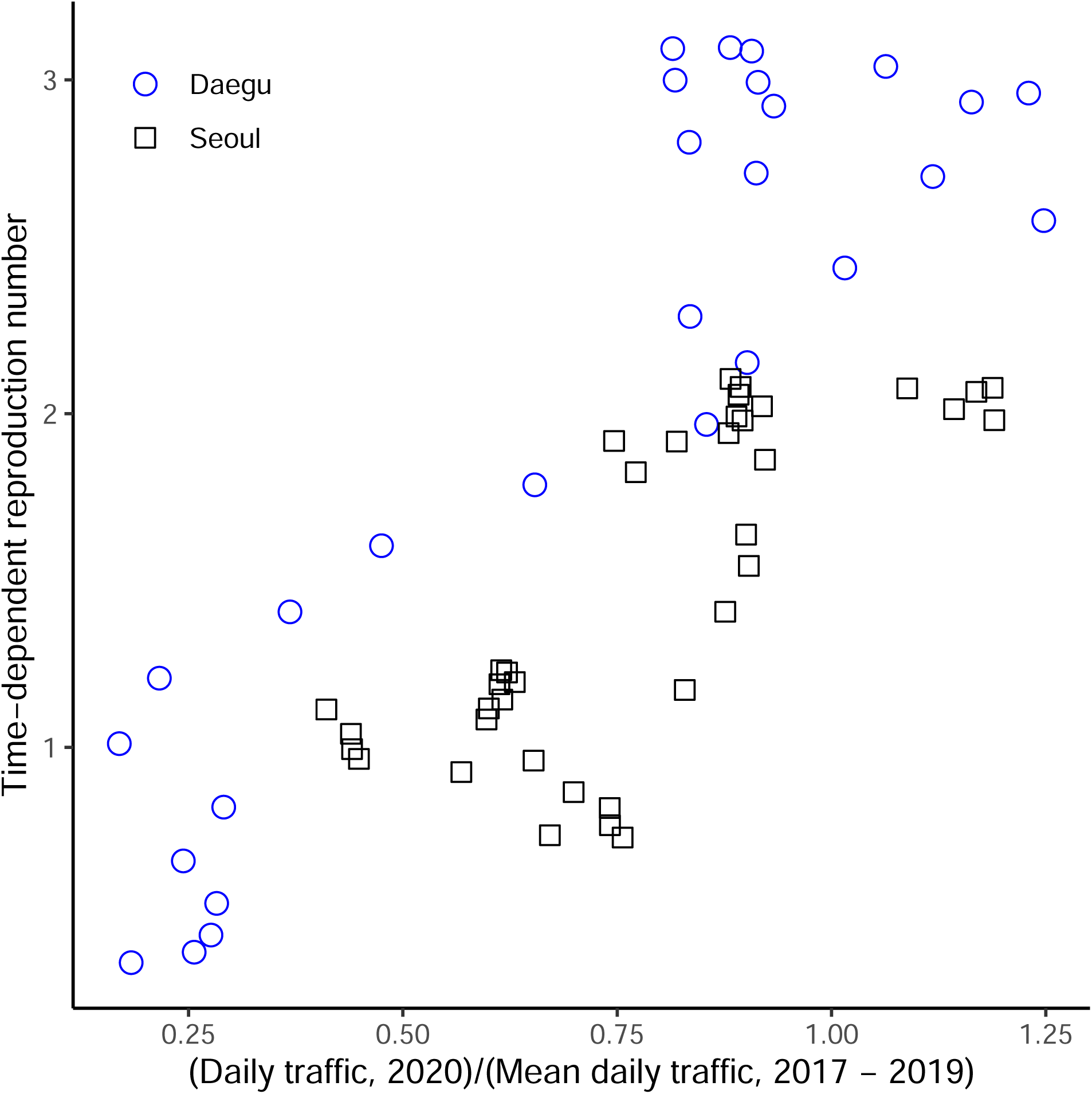
Scatter plot of the normalized traffic volume and the median estimates of. ℛ_*t*_.

**Figure S3:**
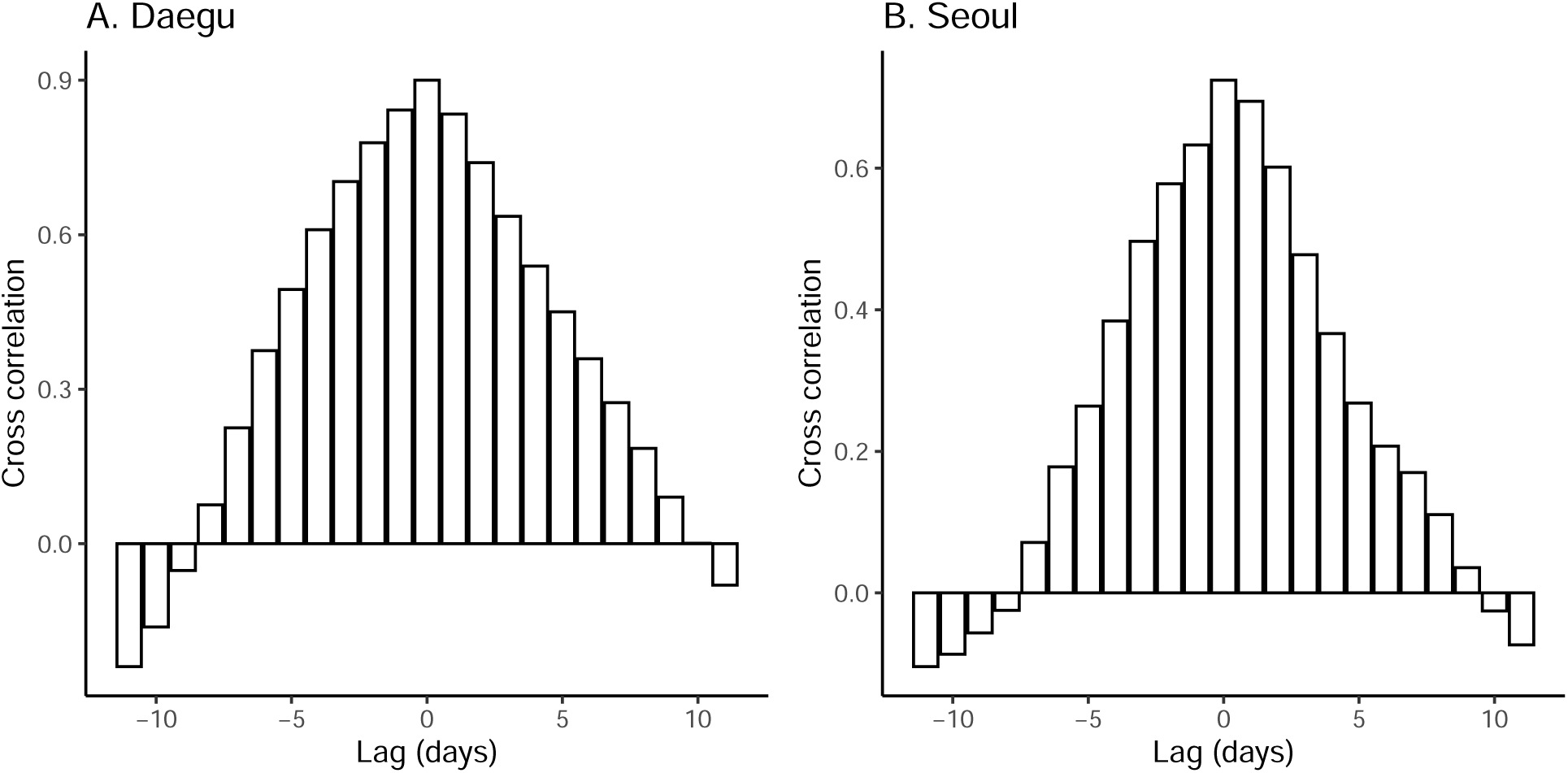
Cross correlation between the normalized traffic volume and the median estimates of. ℛ_*t*_.

**Figure S4:**
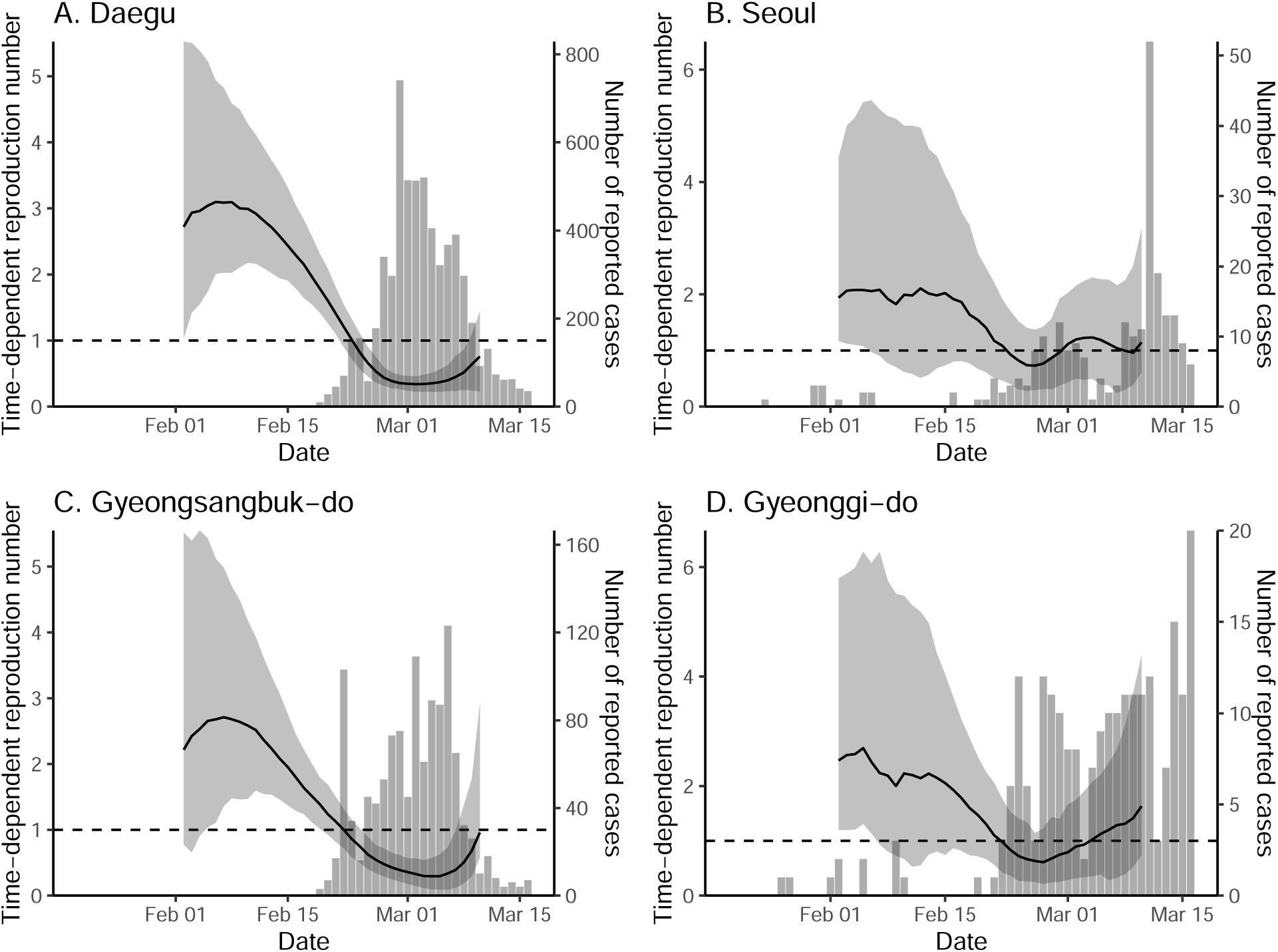
Comparison of time-dependent reproduction number and the daily number of reported cases in Daegu, Seoul, Gyeongsangbuk-do, and Gyeonggi-do.

**Figure S5:**
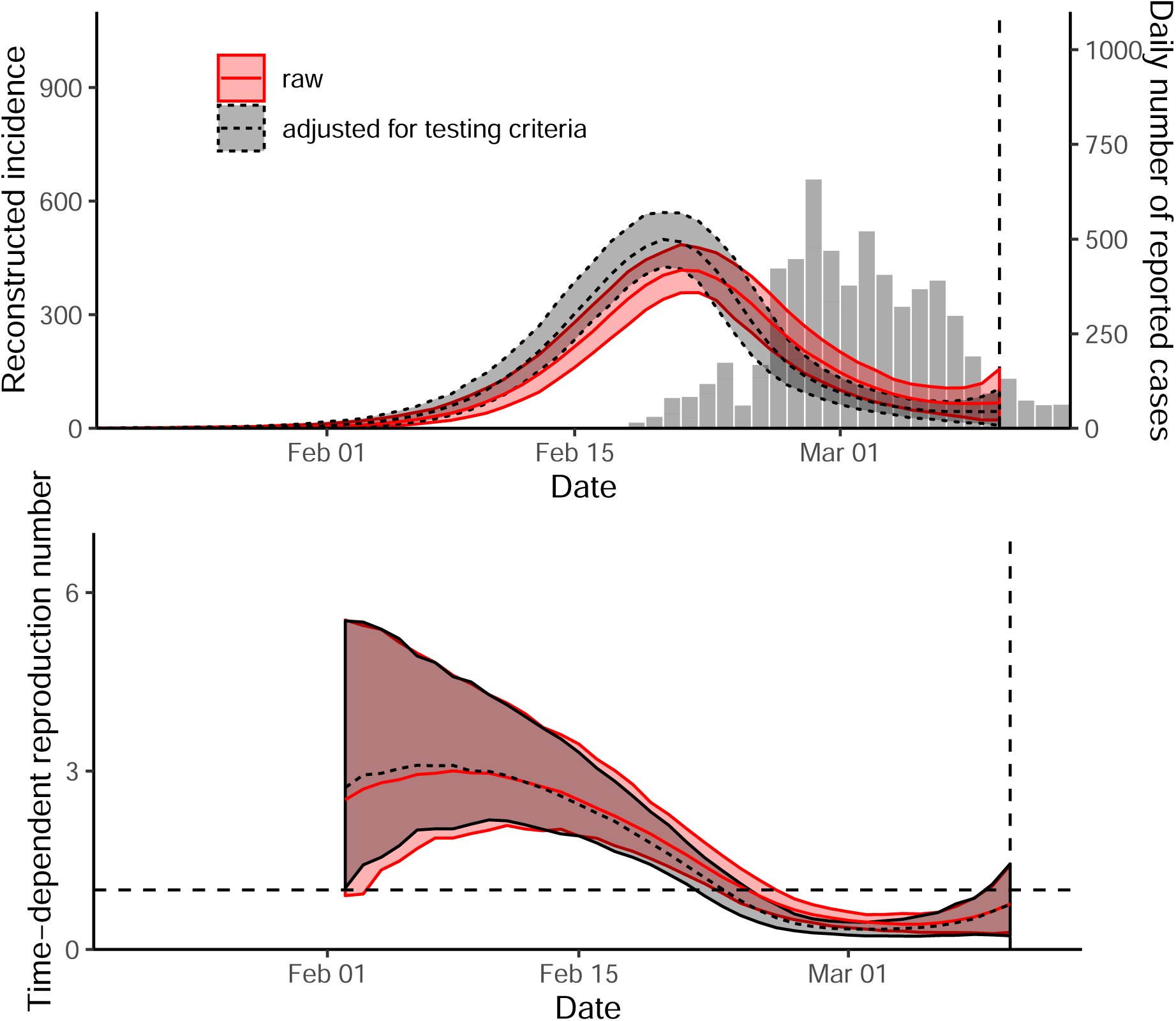
Sensitivity analysis of estimates of. ℛ_*t*_ **in Daegu**.

**Figure S6:**
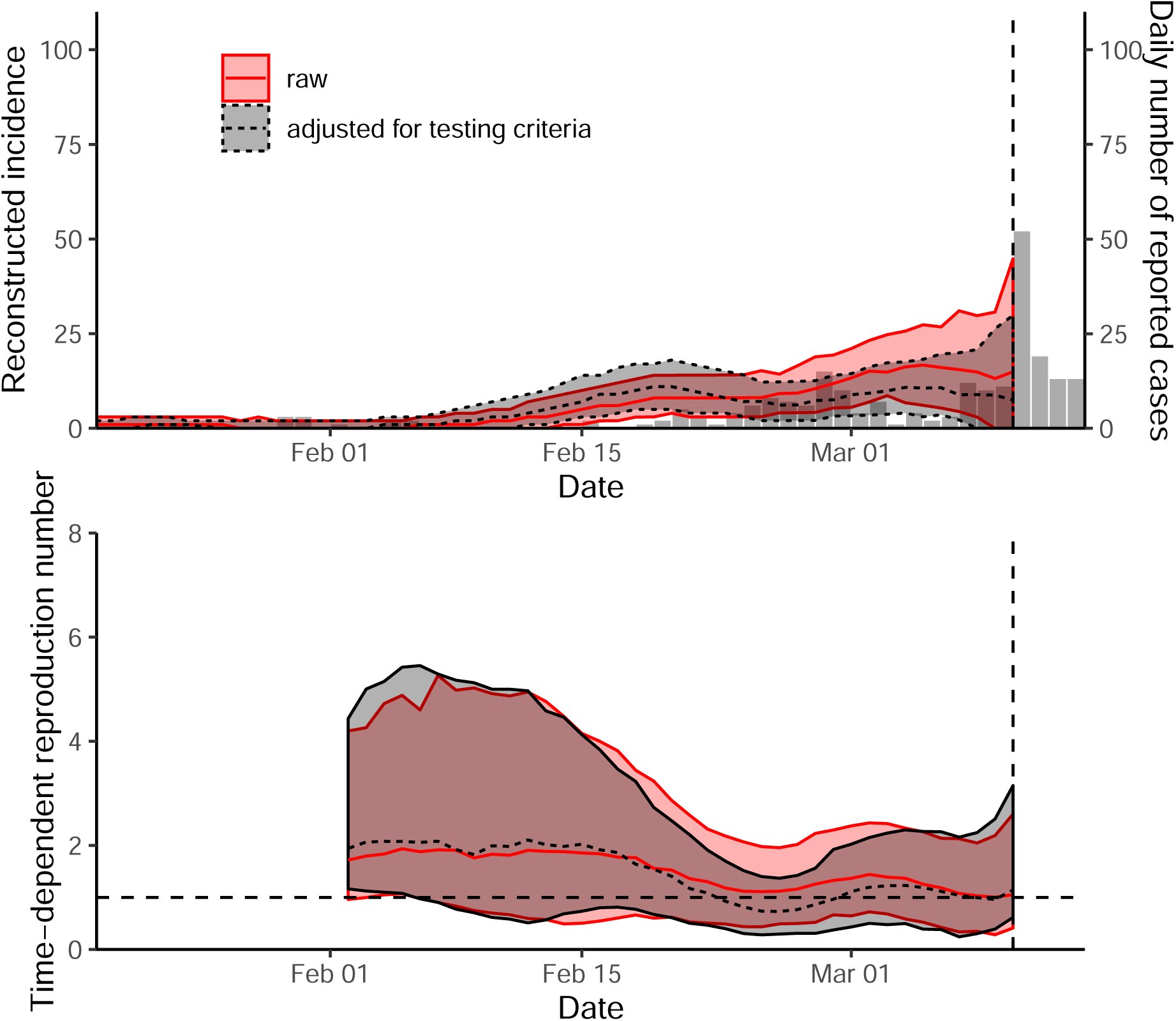
Sensitivity analysis of estimates of. ℛ_*t*_ **in Seoul**.

**Figure S7:**
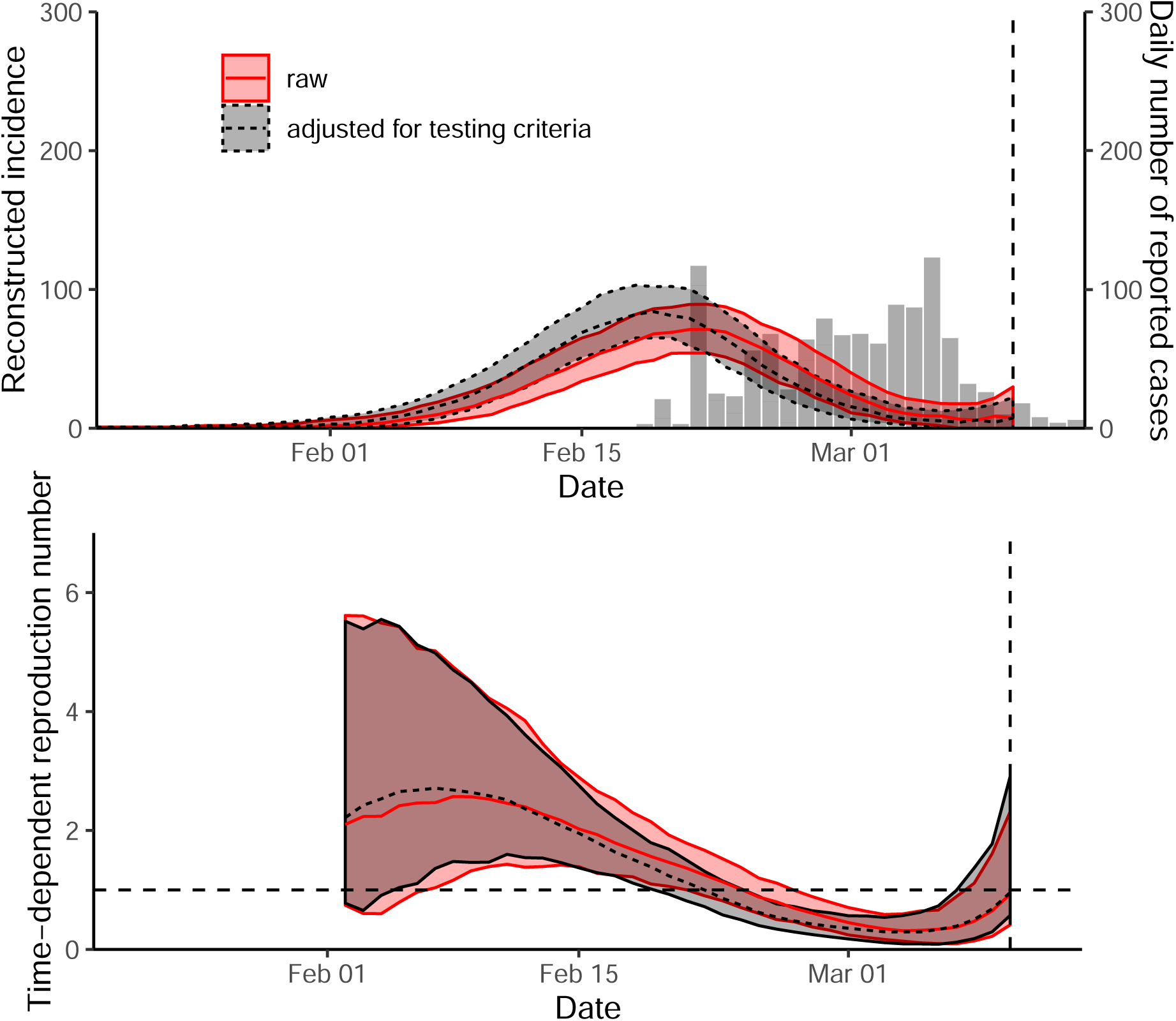
Sensitivity analysis of estimates of. ℛ_*t*_ **in Gyeongsangbuk-do**.

**Figure S8:**
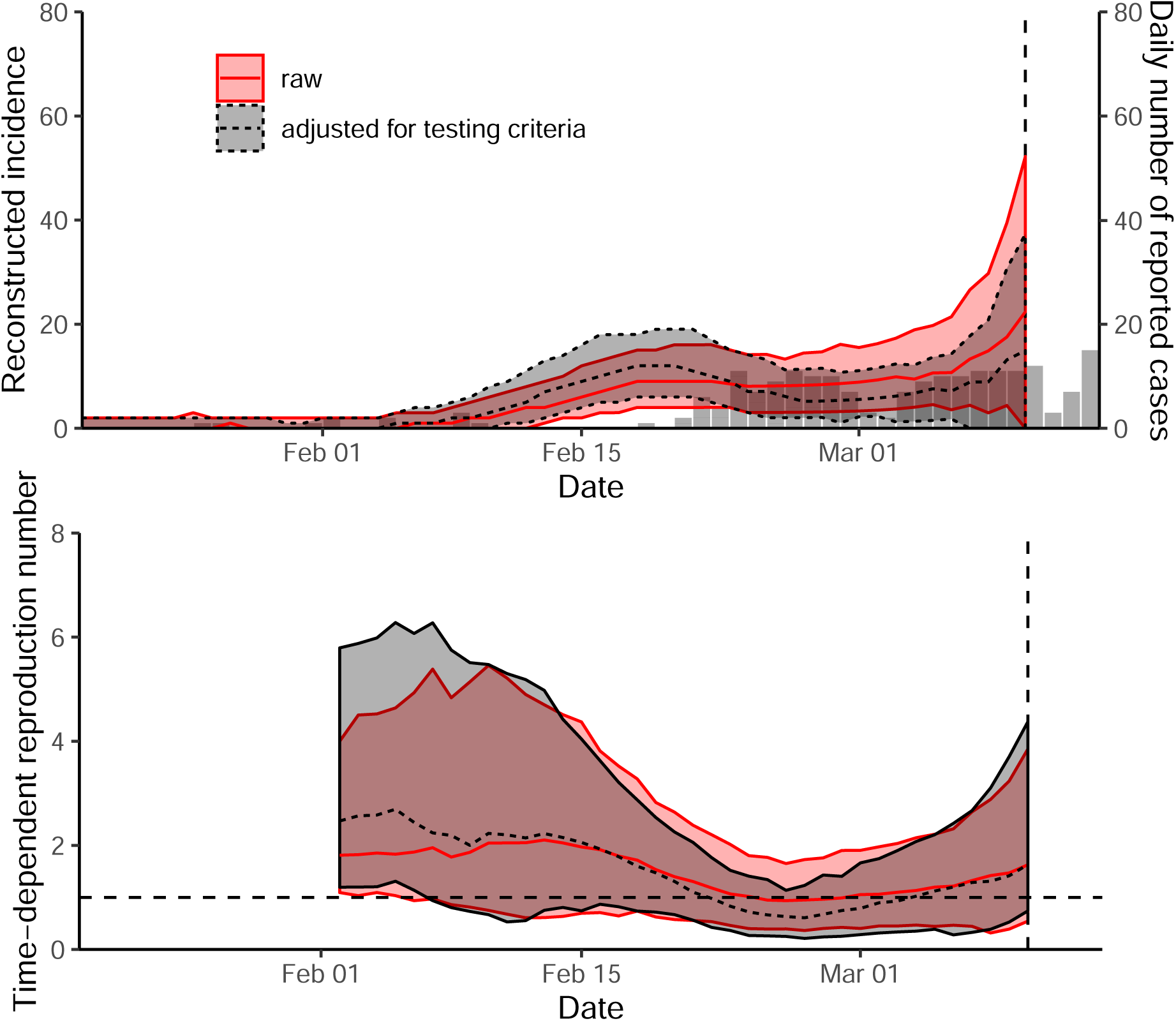
Sensitivity analysis of estimates of. ℛ_*t*_ **in Gyeonggi-do**.

**Figure S9:**
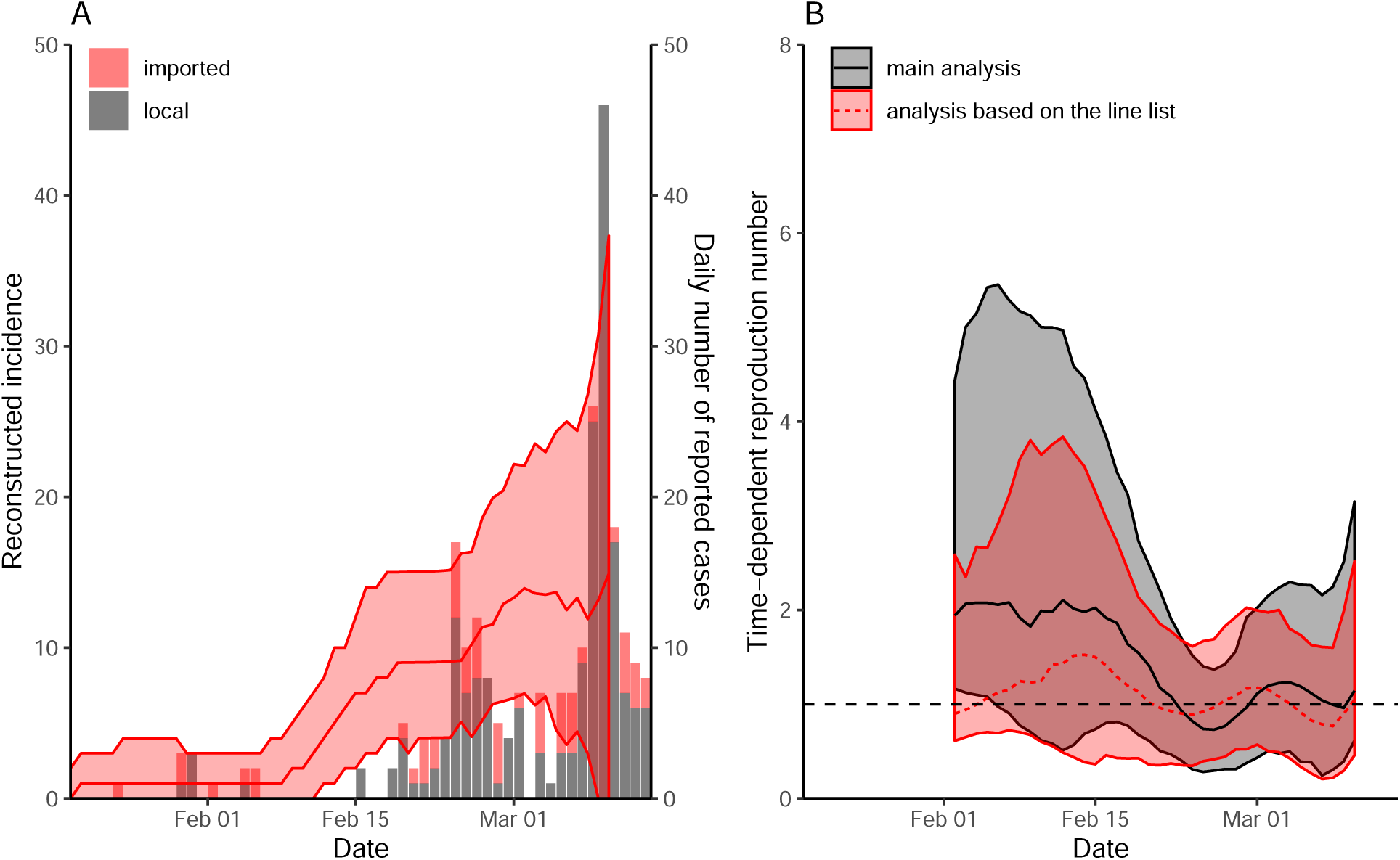
Comparison of time-dependent reproduction number in Seoul using the number of reported cases by the KCDC and the line list provided by the Seoul Metropolitan Government. Using line list, we reconstructed incidence for local 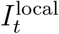 and imported 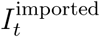 cases separately based on the method described in the main text. Then, we estimated the time-dependent reproduction number via 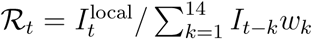, where 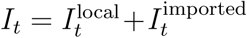 We did not account for changes in testing criteria in this analysis. The line lists were obtained from http://news.seoul.go.kr/welfare/archives/513105.

